# Spatial Clusters and Predictors of Substance-Related Mortality in Iran: A Geographically Weighted Regression Analysis

**DOI:** 10.64898/2025.12.07.25341803

**Authors:** Fatemeh Baberi, Amir Kavousi, Ebbie Kalan, Alexander Hohl, Abbas Rezaianzadeh, Fatemeh Jafari, Yousef Alipour, Samira Dilruba Ali, Seyed Saeed Hashemi Nazari

## Abstract

**Background:** Substance abuse is a critical public health concern worldwide, causing significant social, economic, and health burdens. In Iran this issue is driven by diverse socioeconomic conditions and its position along regional drug trafficking routes. This study aims to explore spatial variations and identify factors contributing to substance-related mortality across provinces and counties.

**Methods:** An ecological study was conducted using a 5-year data from the Iran Legal Medicine Organization (ILMO) in 31 provinces and 424 counties. Descriptive statistics and univariate regression analysis determined the key variables. Geographically Weighted Poisson Regression (GWPR) was employed to assess local impacts and spatial clustering methods were developed to visualize influential variables.

**Results:** A total of 12,632 substance-related deaths were recorded during the study period, with over 90% of cases among men and 36% aged 30–39 years. Mortality rates ranged from 24.6 per million population in Kohgiluyeh and Boyer-Ahmad to 302.13 per million in Hamedan. Spatial clustering of mortality was significant (Moran’s I = 0.015, *p* = 0.014). The GWPR demonstrated superior model performance than ordinary least squares (AIC: 3351 vs. 6764), revealing substantial spatial heterogeneity in risk factors. Percentages of tenant families, older adults, and substance abuse arrests had the strongest positive association. Conversely, substance smuggling arrests, provincial gross income, and distance from key points demonstrated negative associations (p < 0.05).

**Conclusion:** Substance-related mortality in Iran shows significant spatial clustering with geographically varying socioeconomic determinants, indicating need for region-specific prevention and treatment public health strategies rather than uniform national interventions.

## 1 Introduction

Substance use and its adverse effects represent a critical global public health concern, imposing significant social, economic, and health burdens on societies, particularly in low- and middle-income countries (LMICs) (1, 2). Globally, substance use contributes to premature mortality, disability, and health inequities, with approximately 255 million people worldwide using substances and nearly 2.5 million deaths occurring annually due to harmful substance use (7, 8). Substance-related mortality occurs both directly through poisoning and overdose, and indirectly through accidents, violence, and infectious diseases (5, 6), with patterns differing significantly across regions and populations (9). For example, alcohol and substance abuse account for 13.7 deaths per 100,000 person-years in Australia (10) and 4,460 deaths in France from 2011 to 2021 (11). Various risk factors, including economic status, unemployment, education level, access to health services, cultural values, and policies, affect substance use patterns, and these factors vary widely across regions, urban and rural areas, and socioeconomic groups (3, 4)These differences highlight the need to understand the contributing factors and identify areas with high substance-related mortality, especially in LMICs such as Iran, where specific social, cultural, and geographical characteristics affect consumption and its health consequences (1). According to a study conducted in 2017, deaths caused by substance abuse in Iran account for approximately 0.61% of the country’s total annual deaths (2). Iran has a special place in the epidemiology of substance use due to its proximity to Afghanistan, the world’s largest opium producer, and its complex socio-economic conditions (3). This country is not only a major transit route for substances but also exhibits diverse patterns of use, including traditional opium and heroin use as well as stimulants and chemicals (3, 4). This upward trend has raised concerns about the increasing number of substance-related deaths. However, national statistics illustrate substances as a significant cause of preventable deaths (5, 6). More detailed regional analyses reveal substantial variations in death rates and contributing factors across Iranian provinces. (7). Understanding these spatial differences is vital for epidemiological research and health planning, as substance-related mortality is shaped by multiple, geographically variable determinants (1, 8). Therefore, traditional statistical methods cannot accurately reflect the issue’s geographical complexities. In this context, location-specific analytical methods are necessary to identify the impact of local factors. One of these methods is geographically weighted Poisson regression (GWPR), which, unlike classical models that assume the stability of the relationship between variables in space, allows the coefficients to differ in different regions. This capability helps to identify areas with strong or weak effects of factors and, in addition to increasing the accuracy of the model, provides a deeper understanding of the geographical pattern of substance-related mortality (9, 10). The GWPR model is similar to the original geographically weighted regression (GWR) model, but it is designed to handle continuous dependent variables. The use of GWPR in public health research has increased in recent years. It has been applied in studies of chronic diseases, health inequalities, and environmental factors (15). Yet, its use in the field of substance-related mortality, especially in LMICs such as Iran, is limited. Therefore, this study aims to determine whether GWPR is an effective tool for analyzing the status of substance use and its influencing factors in different regions of Iran. Can GWPR better capture spatial variability compared to traditional regression models and quantify the extent of differences between these approaches? This will facilitate the identification of high-risk areas and local predisposing factors, thereby supporting the design of targeted interventions, such as social protection programs, tailored to vulnerable communities.

## 2 Material and Methods

### 2-1 Study setting

Iran is located in Southwest Asia, covering 1,648,195 km^2^ and ranks 5th in Asia in land coverage, with a great geographical diversity, and has a population of more than 85 million in 31 provinces.

Afghanistan and Pakistan border Iran from the east, Turkey and Iraq from the west, Azerbaijan, Armenia, and Turkmenistan from the north, and are also bordered by the Caspian Sea from the north and the Persian Gulf and the Sea of Oman from the south. Iran is between 25 and 40 degrees north latitude and 44 and 64 degrees east longitude(11). This was an ecological county-level study conducted at 424 counties of Iran.

### 2-2 Population

Generally, 15,304 deaths had been registered in the Iran Legal Medicine Organization (ILMO) due to substance abuse from 2014 to 2018. Data were collected for 12632 of the 15304 individuals using abstracting tools, including socio-demographic characteristics and autopsy results.

### 2-3 Variables of the study

The outcome variable used for this study was the number of deaths due to substance abuse. The substances examined in this study included codeine, diphenoxylate, methadone, morphine, tramadol, amphetamine, methamphetamine, and benzodiazepines. Details of these substances have been published in a previous study of this national project(12). Independent variables of the study include population of 20-40 years old, population of male group, illiteracy, marriage ratio, non-Iranian nationality, urbanization, distance from substance transit zone, GDP, number of substance treatment centers, economic participation, tenant, house hold members, breadwinner women, population of up to 60 years, population of arrived, immigrants, substance arrest & seizure, Gini index, number of individuals who arrested due to substance consume, number of individuals who arrested due to substance transition. Variables were selected based on evidence from a literature review, which was confirmed by the opinions of ILMO professionals and Drug Enforcement Administration (DEA) experts. A detailed table and explanation to clarify how field observations and expert interviews guided the operationalization and refinement of the variables are provided in Table S1.

### 2-4 Data management and analysis

We used 3 data sources in this study: we received the substance-related deaths data from the ILMO, province and county level maps from the National Cartographic Center of Iran (NCC), and we extracted the independent variables of our study from the census data, which were recorded in the Statistical Center of Iran. At the first step, descriptive analysis was conducted on the ILMO. Literature review, experts’ opinions, and the reports of the DEA help us to regulate and adjust the desired variables of study from the raw census. During the data collection stage for deceased individuals at ILMO, to ensure the accuracy of the process, the doctor in charge of the autopsy room was selected as the provincial project manager. They were responsible for training other colleagues to complete the forms and check the accuracy of the information. The provincial official, besides the monitoring of the form completion, rechecked the correctness of the information and confirmed it.

In this ecological study, we first performed a univariate regression to identify the significance of the variables at a p-value of 0.2, and then we modeled the significant ones. We used Excel for data management and Stata 17 for primary analysis.

### 2-5 Spatial analysis

We performed spatial autocorrelation analysis to assess whether substance-related mortality rates and their predictors show spatial clustering. Global indices (Global Moran’s I(13), Getis-Ord General(14)) were used to examine overall spatial autocorrelation across the counties of Iran, while local indices (Local Moran’s I(15) and Getis G*(14)) were applied to find specific clusters and hot spots. Using Local Moran’s, I there will be 4 probable types of results including High–High (H-H) clusters, which represent counties with high mortality surrounded by other high-rate counties (“hot spots”), while Low–Low (L-L) clusters indicate low-rate areas grouped together (“cold spots”). Spatial outliers include High–Low (H-L), counties in which high-rate counties are adjacent to low-rate areas, and Low–High (L-H), in which low-rate counties are adjacent to high-rate areas. These analyses were done using ArcGIS 10.7. The Geographically Weighted Poisson Regression (GWPR) method was applied to investigate the variation of the determinant at the county level with the GWR software(16). The results of this analysis were the risk factors affecting the substance-related deaths in each area, and then we entered the results of the analysis with GWR software into ArcGIS 10.7 software and presented the maps for all variables. In other words, we conduct a spatial regression model for each county, using the specified predictors.

### 2-6 Geographically Weighted Regression

In order to show how geography can affect the substance-related deaths in the counties of Iran, we conduct a GWPR analysis. An Ordinary Least Squares (OLS) was also conducted to confirm that when studying factors dependent on geography, it would not be reasonable to adopt a single regression model for the entire study area.

Using the GWPR method, we investigated the status of 19 variables across 424 counties of the country. The selected model was the Poisson. We used an adaptive kernel with a Gaussian function to spatial weights determination. The golden selection option of the software was applied to choose the best bandwidth(17) (Limit: Min: 32, Max: 422, Best bandwidth: 46), which resulted in the lowest AIC value for the model. Finally, we visualized the variation of each factor throughout the whole country.

## 3 Results

### 3-1 Socio-Demographic and other Characteristics

A total of 12,632 deaths caused by substance abuse were investigated from 2014 to 2018. Over 90% of the deceased were Iranian, Afghans comprising the highest frequency at 1.53%, among other nationalities. Approximately 5% of the deceased had an unknown status. According to the information obtained, 649 (5.13%) samples of the deceased had no identifiable identity. Of those, 1960 individuals (15.52%) were reported to have died due to overdose. Only 18.43% of the deceased had a history of arrest, and 4.3% had a history of suicide. Table 1 presents detailed characteristics of the study population.

**Table 1:** Demographic characteristics of study sample (N=12632)

| **Variables** | **Number** | **%** |
| --- | --- | --- |
| **Gender** |  |  |
| Male | 11403 | 90.27 |
| Female | 1229 | 9.73 |
| **Age group** |  |  |
| 0-9 | 480 | 3.79 |
| 10-19 | 522 | 4.1 |
| 20-29 | 3030 | 23.99 |
| 30-39 | 4593 | 36.36 |
| 40-49 | 2286 | 18.09 |
| 50-59 | 1149 | 9.09 |
| 60-69 | 384 | 3.04 |
| 70-79 | 136 | 1.08 |
| 80-100 | 52 | 0.41 |
| **Marital Status** |  |  |
| Single | 4324 | 34.23 |
| Married | 4578 | 36.24 |
| Divorced | 1292 | 10.22 |
| Widow | 124 | 0.98 |
| Unknown | 2314 | 18.31 |
| **Education** |  |  |
| Low | 9792 | 77.51 |
| Medium | 692 | 5.47 |
| High | 62 | 0.49 |
| Unknown | 2086 | 16.51 |
| **Living** |  |  |
| Alone | 2406 | 19.05 |
| Not alone | 8547 | 67.65 |
| Unknown | 1679 | 13.28 |
| **Nationality** |  |  |
| Iranian | 11676 | 92.43 |
| Afghan | 194 | 1.53 |
| Iraqi | 13 | 0.1 |
| Other | 109 | 0.86 |
| Unknown | 640 | 5.06 |
| **Treatment condition** |  |  |
| Under-treatment | 2067 | 16.36 |
| Not under treatment | 6996 | 55.38 |
| Unknown | 3569 | 28.25 |
| **Overdose history** |  |  |
| Positive | 1960 | 15.52 |
| Negative | 7134 | 56.47 |
| Unknown | 3538 | 28.01 |
| **Arrestment history** |  |  |
| Positive | 2328 | 18.43 |
| Negative | 7476 | 59.18 |
| Unknown | 2828 | 22.39 |
| **Suicide history** |  |  |
| Positive | 543 | 4.3 |
| Negative | 9310 | 73.7 |
| Unknown | 2779 | 21.99 |
| **Identity** |  |  |
| Known | 11983 | 94.86 |
| Unknown | 649 | 5.13 |

### 3-2 Substance-related mortality rates

The incidence rate of substance-related mortality in the provinces and counties of Iran per 1 million population during 2014-2018 is depicted in Figure 1. The highest incidence rate is in Hamedan Province, followed by Zanjan. The lowest rates are in Kohgiluyeh and Boyer-Ahmad Province, and Tehran. Results from Khudafarin, Heris, Bostanabad, and Ajabshir (East Azerbaijan); Khorramdarreh (Zanjan); Aradan (Semnan); Razan and Hamedan (Hamedan Province); Khorramabad (Lorestan); Zahedan (Sistan and Baluchestan); and Shahin (Isfahan) shows that these ten counties report the highest death rates nationally. Incidence rates for all provinces and counties are provided in Table S2 in the supplementary.

**Fig 1:**
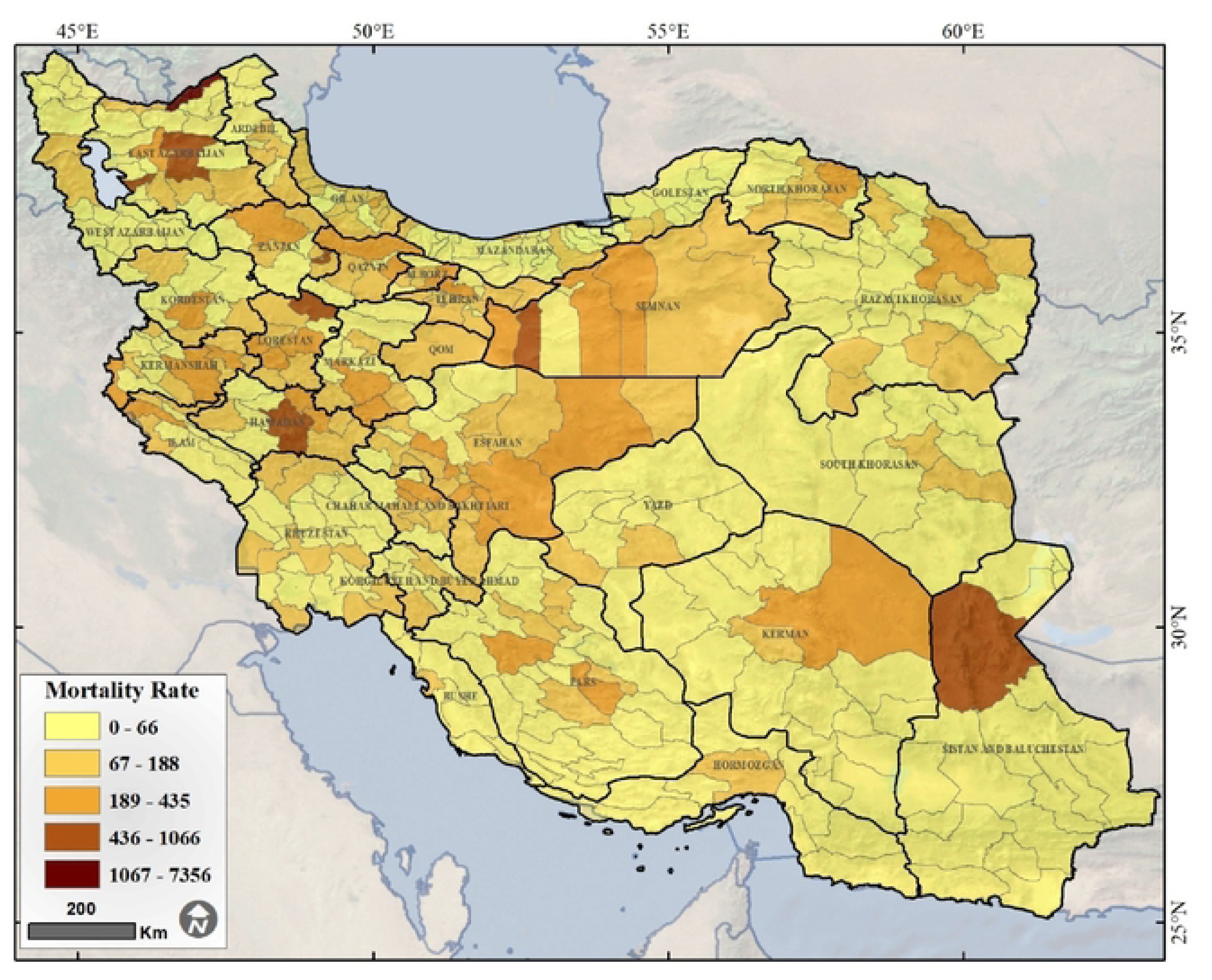
substance-related mortality in the provinces and counties of Iran per million

### 3-3 Spatial Autocorrelation of Substance-related mortality rates in Iran

The Global Moran’s index [P=0.014, I=0.015] indicates that the distribution of deaths in Iranian counties is not random and presents positive spatial autocorrelation (clustering). To assess the presence or absence of high or low spatial clustering, we used the Getis-Ord General. According to the value of this index, G = 0.056 and the significance level of the p-value = 0.299, the results indicate that the distribution of counties with high and low values of deaths caused by substances together was random. Figures S1 and Figure S2 in the supplementary show the Global Moran’s I and Getis-Ord General results, respectively.

#### 3-3-1 Cluster and Outlier of Substance-related mortality

Local Moran’s I statistics showed the spatial clustering pattern of substance-related mortality rates in Iran’s counties. This index allowed us to determine in which high or low value of mortality clusters were observed, and to identify the areas whose values are significantly different from their neighbors, named outliers. This analysis revealed that the H-H clusters were mainly concentrated in East Azerbaijan province, located in the northwest of Iran. In contrast, L-L clusters were scattered in different regions of the country, with most of these clusters located in the south and southeast of Iran, especially in the counties of Sistan and Baluchestan province and the southern counties of Kerman province. Many Clusters were also observed in the provinces of Yazd, Khuzestan, southern Fars, Mazandaran, and Golestan. Outliers were spread in different regions. Details of this analysis are reported in Table S3 in the supplementary. Figure 2 illustrates the findings from the Local Moran’s I analysis.

**Fig 2:**
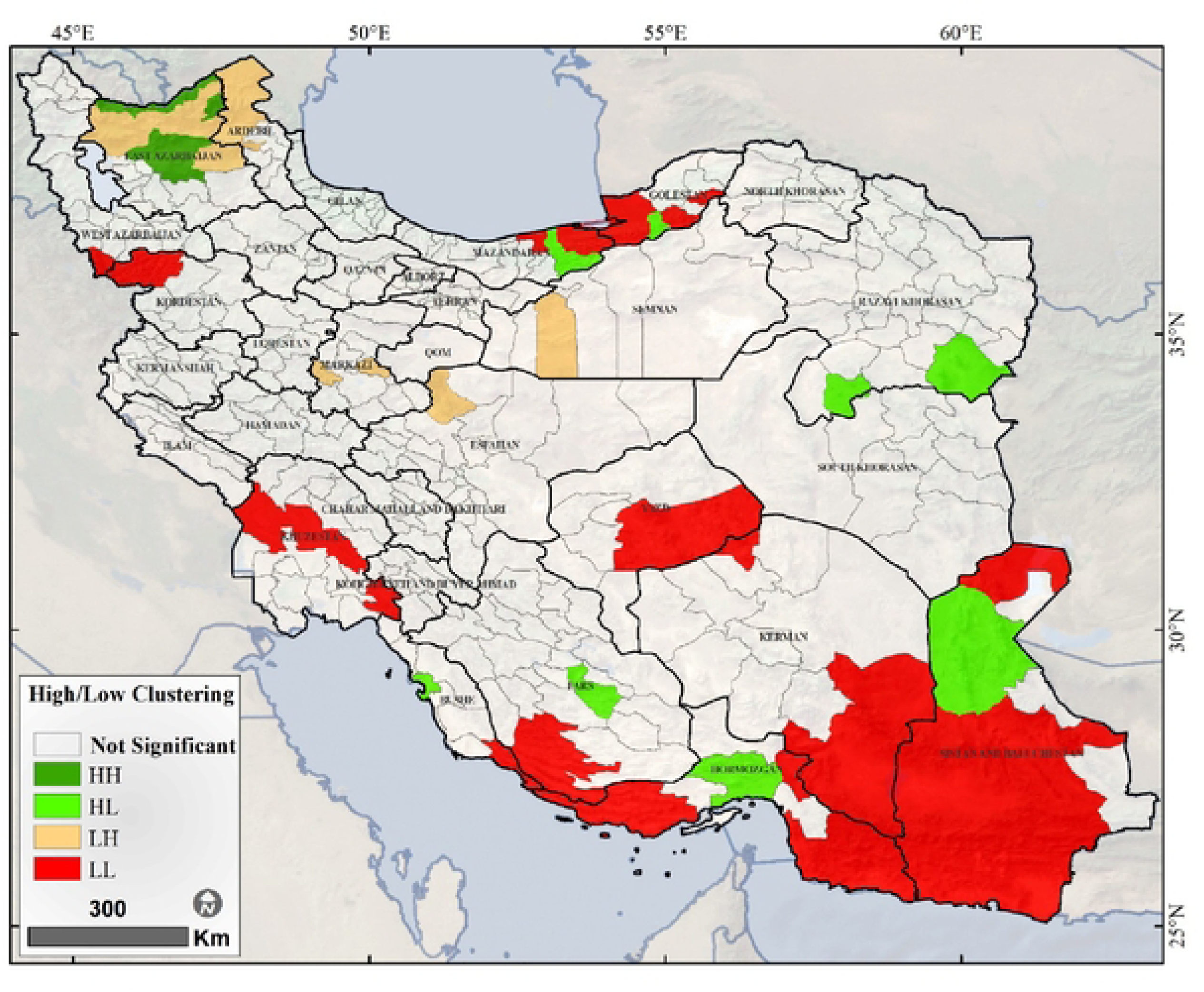
Cluster and Outlier of Substance-related mortality rates in Iran (Local Moran’s I)

Table S3: Cluster and Outlier of Substance-related mortality rates in Iran (Local Moran’s I)

#### 3-3-2 Hot spot and cold spot of substance-related mortality

Based on Getis-Ord Gi* statistic, no cold spots were found at the county level. Identified hot-spots were located in northwest of Iran in East Azerbaijan and Ardabil provinces. Figure 3 depicts the results of Getis-Ord Gi* analysis.

**Fig 3:**
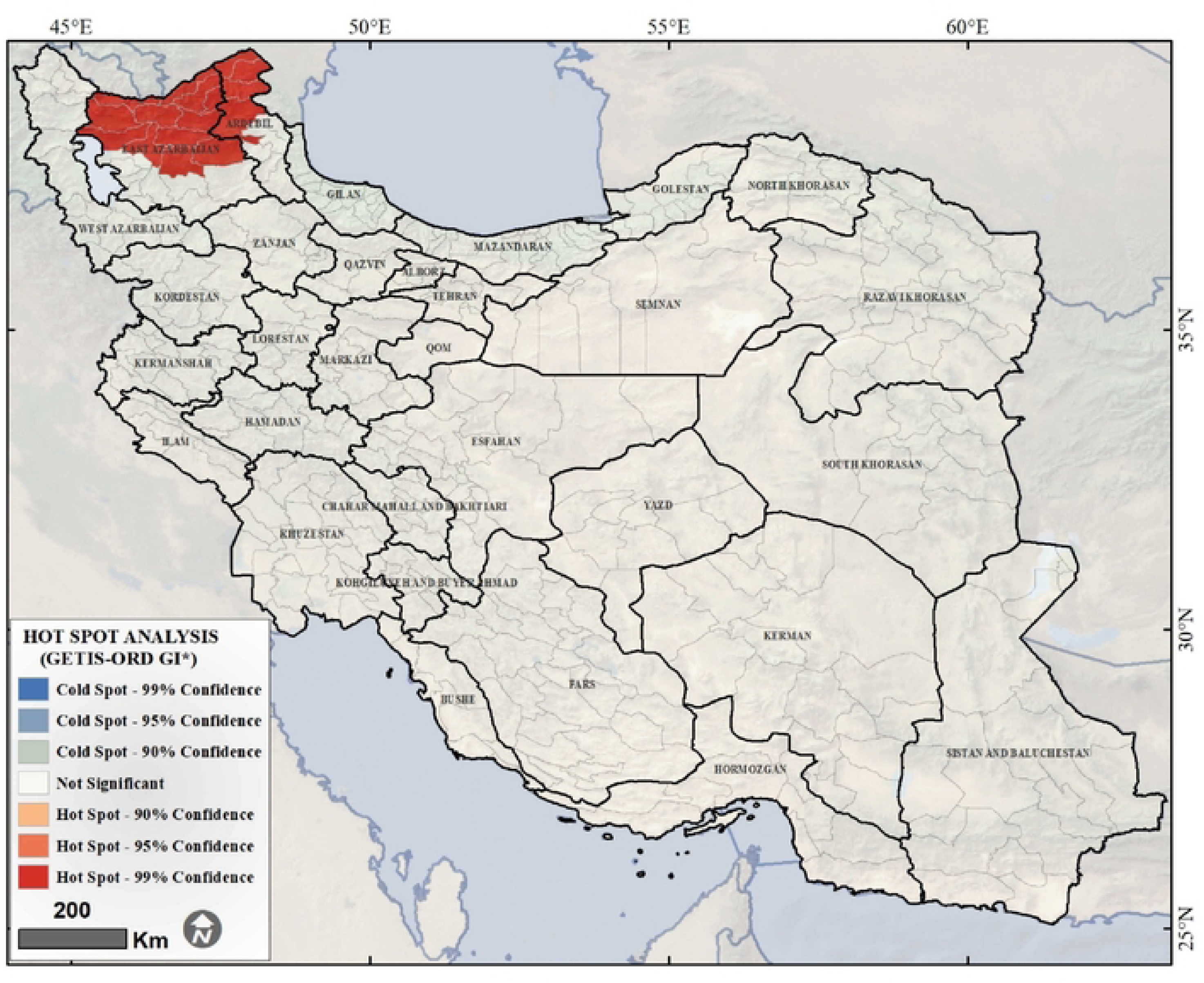
Hot-spot Analysis of Substance-related mortality rates in Iran (Getis-Ord Gi*)

### 3-4 Ordinary least squares analysis for factors associated with substance-related mortalities

All included variables in the OLS model suggested a significant relationship with substance-related mortality, except the average number of household members, marriage rate, male population, age group 20-40 and urban residents (Table 2).

**Table 2:** General regression results performed for study variables

| Variable | $\beta$ (Estimate) | SE | Z | Exp($\beta$) [95% CI] | p-value |
| --- | --- | --- | --- | --- | --- |
| % Tenant families | 0.59 | 0.02 | 26.86 | 1.80 [1.73–1.87] | < 0.001 |
| % Aged ≥ 60 years | 0.26 | 0.02 | 10.76 | 1.30 [1.25–1.36] | < 0.001 |
| % Substance abuse arrests | 0.25 | 0.03 | 9.39 | 1.29 [1.23–1.36] | < 0.001 |
| Illiteracy rate | 0.13 | 0.02 | 5.78 | 1.14 [1.09–1.19] | < 0.001 |
| % Immigrants | 0.12 | 0.01 | 8.98 | 1.13 [1.10–1.17] | < 0.001 |
| Gini coefficient | 0.11 | 0.01 | 9.40 | 1.12 [1.09–1.16] | < 0.001 |
| % Female-headed households | 0.06 | 0.02 | 2.85 | 1.06 [1.02–1.10] | 0.002 |
| Addiction treatment centers | 0.04 | 0.00 | 8.53 | 1.04 [1.03–1.05] | < 0.001 |
| Economic participation | −0.05 | 0.02 | −2.18 | 0.95 [0.91–0.99] | 0.030 |
| % Non-Iranian nationality | −0.04 | 0.01 | −3.74 | 0.96 [0.94–0.98] | < 0.001 |
| Discovered materials | −0.07 | 0.02 | −3.64 | 0.93 [0.89–0.97] | < 0.001 |
| Distance from key points | −0.11 | 0.02 | −5.08 | 0.89 [0.85–0.93] | < 0.001 |
| Provincial gross income | −0.26 | 0.02 | −16.74 | 0.77 [0.74–0.80] | < 0.001 |
| % Substance smuggling arrests | −0.36 | 0.04 | −8.13 | 0.70 [0.64–0.77] | < 0.001 |
| % Urban residents | 0.03 | 0.03 | 1.14 | 1.03 [0.98–1.08] | 0.240 |
| Marriage rate | 0.02 | 0.02 | 1.19 | 1.03 [0.98–1.07] | 0.220 |
| % Aged 20–40 years | 0.02 | 0.02 | 1.20 | 1.02 [0.99–1.05] | 0.220 |
| % Male | −0.03 | 0.03 | −1.11 | 0.97 [0.91–1.03] | 0.260 |
| Household members | −0.00 | 0.01 | −0.33 | 0.74 [0.68–0.81] | 0.996 |

### 3-5 Spatial analysis of predictors of substance-related mortality

The effect of geographic factors on substance-related deaths in Iranian counties was measured using a GWPR model. This model utilized an adaptive kernel with a Gaussian function, and the bandwidth was selected using the golden option, which set the minimum and maximum points of the search based on the lowest AIC value. Comparing the fitted GWPR model to the OLS, the AIC decreased from 6764 to 3351. The selected model accurately represented the relationship between substance-related mortality and the variables examined in this study.

### 3-6 The Goodness of Fit (GOF) of the GWPR Model

#### 3-6-1 GOF of model based on Yhat’s

To assess the model fit, we mapped the predicted substance-related mortality (Yhat’s) using the GWPR model (Figure 4). The predicted patterns nearly match the observed mortality rates across the country as reported in Figure 1, declaring that the GWPR model precisely captured the spatial distribution of the substance-related deaths.

**Fig 4:**
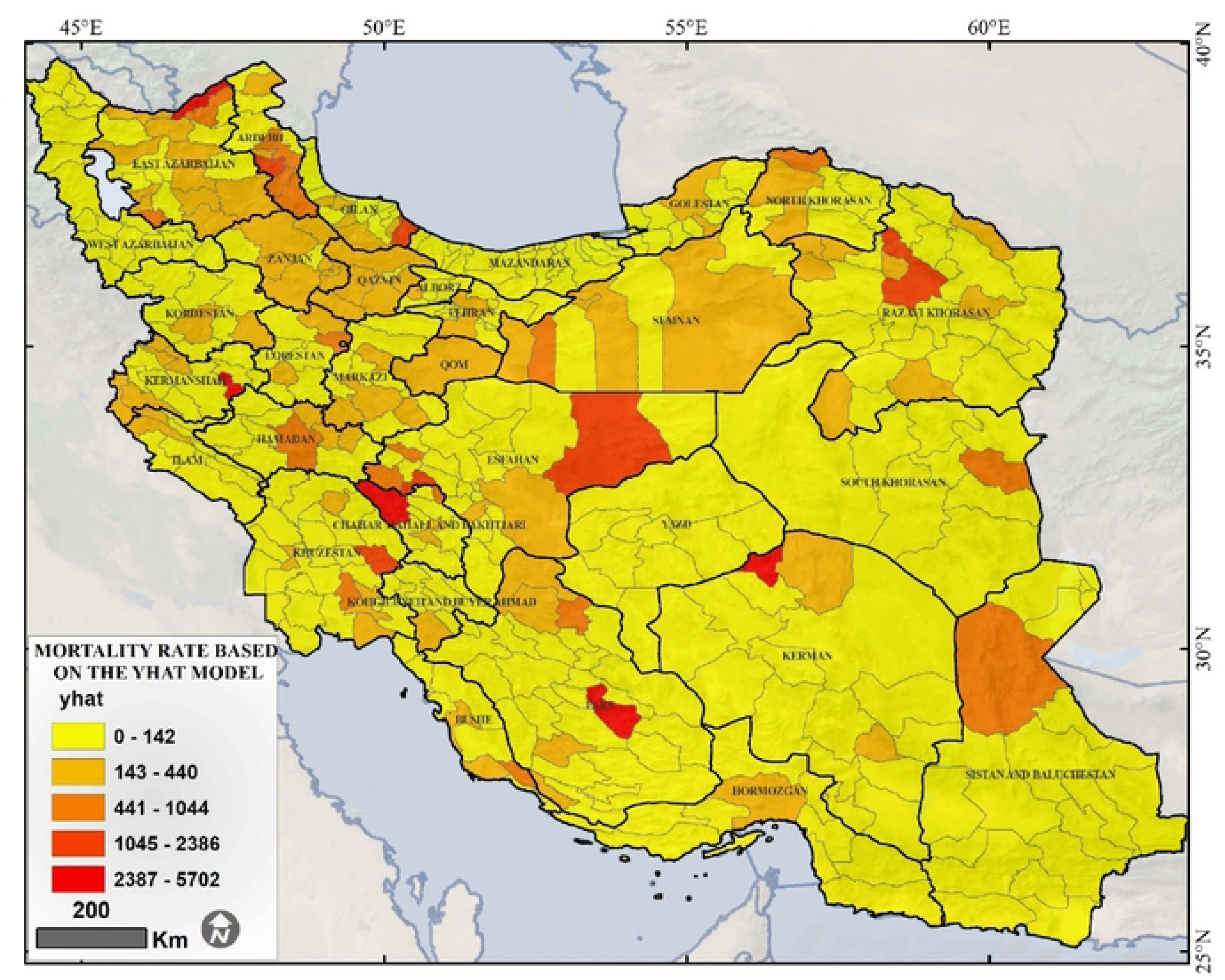
substance-related mortality rate per million based on Yhat’s

**Fig 5:**
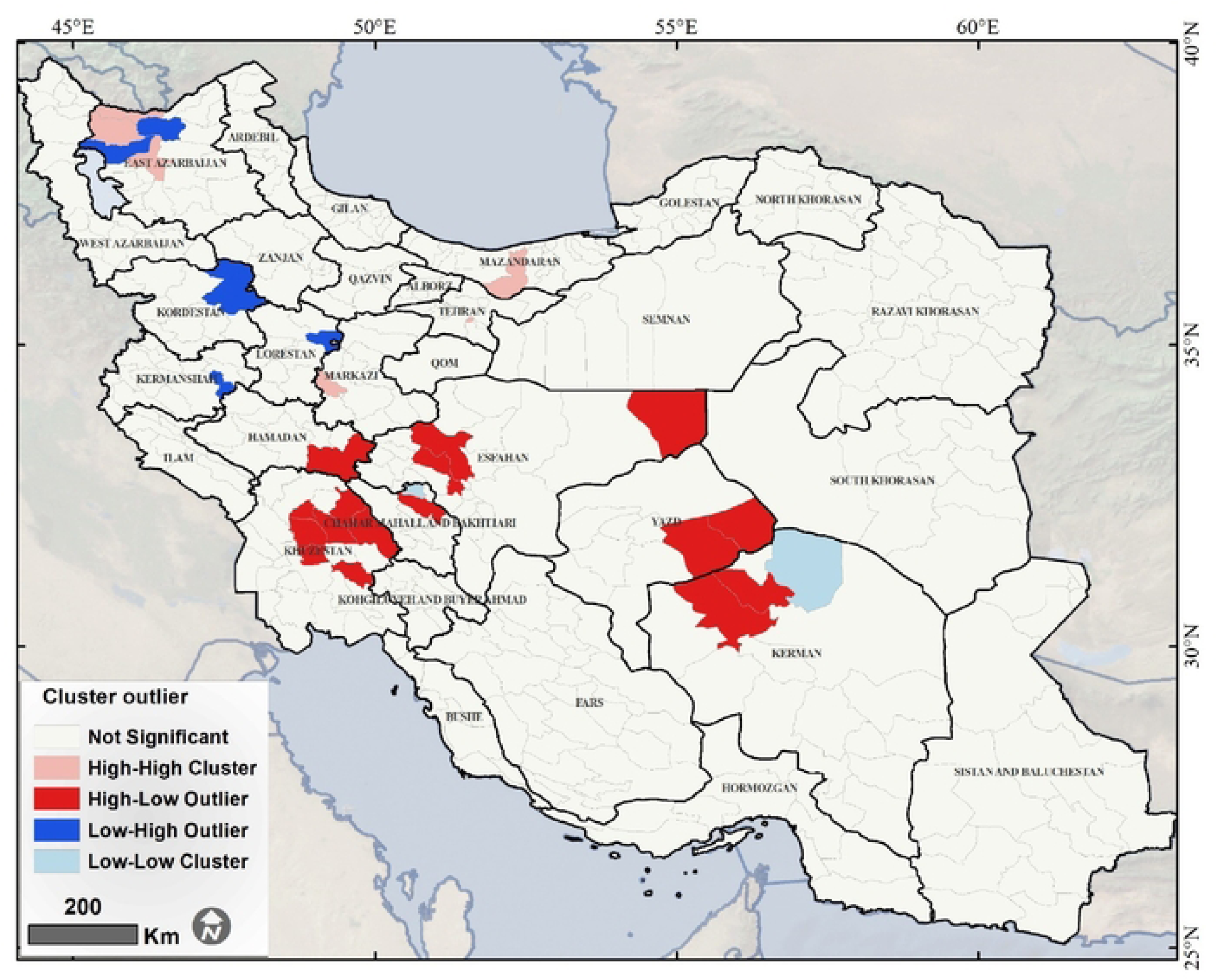
Distribution of residuals of GWPR model

#### 3-6-2 GOF of model based on Residuals

we examined the spatial distribution of residuals and identify significant clusters to evaluate the fitting of the model. To do this, we applied the Local Moran’s I test. Counties that are classified as High–High (H-H) indicate regions where both the model residuals and neighboring counties were high, suggesting weak model prediction in these areas. Counties in East Azerbaijan, Mazandaran, Tehran, and Markazi provinces formed H-H clusters in the residuals analysis, indicating areas where the GWPR model under-or over-predicted substance-related mortality.

## 4 Discussion

This study provides the first comprehensive spatial analysis of substance-related mortality across Iran, revealing that mortality patterns from 2014 to 2018 were not random. The findings demonstrate significant spatial autocorrelation (Global Moran’s I=0.015, p=0.014), with a distinct high-mortality cluster and hot spot identified in the northwestern provinces, particularly in East Azerbaijan. Demographically, the burden of mortality was overwhelmingly borne by males (90.3%), with the 30-39 age group being the most affected (36.4%). Crucially, this analysis demonstrates that a geographically weighted regression model was substantially superior to a standard OLS model, confirming that the factors associated with substance-related mortality are not uniform and vary significantly by geographic location. These results underscore the critical need for spatially-targeted, regional public health interventions rather than a single, national-level strategy.

Consistent with our findings, previous studies(12, 18) have reported that most deaths occur among men in the 30-39 age group. This pattern may reflect gender differences in social mobility, as men typically experience greater freedom in family and social relationships across most societies. In Iranian culture specifically, women’s relationships are subject to closer family oversight than men’s, resulting in men having greater accessibility and exposure to risk factors(19). Our findings regarding marital status, however, present a different pattern. While some Iranian studies have reported higher mortality rates among unmarried individuals(2, 19), our results align with those of another study in showing a different trend(20). The elevated risk among married individuals in our study may reflect the compounding pressures of family responsibilities and unfavorable economic conditions that disproportionately affect this group. These findings highlight the multifaceted nature of mortality risk factors and suggest that further research is needed to fully elucidate the mechanisms underlying these demographic and social patterns, particularly regarding marital status and economic pressures, which have become even more pronounced following the COVID-19 pandemic(21, 22).

Our findings showed that high-risk areas are mainly concentrated in East Azerbaijan province, located in the northwest of Iran. This geographic pattern aligns with known drug trafficking routes to the EU, as Iran borders Afghanistan, one of the world’s primary source of opium(23). The Afghanistan-Pakistan corridor through eastern Iran to Turkey in the northwest serves as a major heroin trafficking route connecting Afghanistan to Europe(24). Beyond its position along this trafficking corridor, East Azerbaijan has experienced rapid urbanization and high population density, leading to the expansion of informal settlements and slums, which are associated with increased substance use(25). The cultural and behavioral context of the region further compounds this risk; previous studies from East Azerbaijan have reported high prevalence of opium consumption as a risk factor for several health outcomes, including cancer(26), suggesting deeply embedded patterns of substance use that contribute to elevated mortality. In contrast, a study in the United States identified high-risk areas in the northeast of the country(27), indicating that geographic patterns of substance-related mortality vary across different national contexts.

While our results showed low-risk areas in the south and southeast of Iran, this pattern warrants careful interpretation and may not solely reflect lower levels of substance-related mortality. Rather, weaker death registration systems, under-reporting, and misclassification of causes of death in these less-resourced regions could contribute to the observed lower rates. A recent study on the completeness of death registration in Iran demonstrates that although national registration completeness improved from approximately 80% in 2006 to 94% in 2021, substantial provincial disparities persist. Provinces in more remote or under-resourced areas often have weaker mortality registration systems, which may lead to systematic underestimation of substance-related mortality in these apparently low-risk regions (28).

According to the OLS model, as with our findings, Meng and colleagues(29) concluded that the lower the socioeconomic status and education, the higher the death rate from substance use. A number of factors may drive higher levels of opioid overdose mortality among less educated people. One important factor is that less educated people are mostly unemployed or employed in low-skilled sectors. As a result, they face an increased risk of depression or anxiety, workplace injuries, and chronic illnesses (30, 31), leading to a greater likelihood of prescription and addiction to painkillers or opioids. Previous findings also confirm that people living below the poverty line are more likely to die from substance overdoses than those living above the line (32, 33). In our study, the Gini coefficient and provincial gross income was also associated with mortality of substance use. A study also found that substance overdose deaths were higher in counties with higher income inequality (34). Other factors like percentage of substance abuse arrests, percentage of substance smuggling arrests, number of addiction treatment centers, distance of the county from the key points, percentage of immigrants, percentage of people over 60 years old, percentage of female household heads, discovered materials, and percentage of tenant families that we have examined in the OLS model were not seen in other studies. It is therefore suggested that more studies be conducted in this area to enable more comparisons.

### Future direction

Given that the present study was conducted ecologically and there was considerable variation in the risk of death across different parts of the country, it is recommended that future studies examine the risk factors in each province based on the endemic risk factors of that region, using individual data. This approach can help inform decision-making and region-specific interventions to control the conditions better and reduce the mortality rate.

### Strengths and Limitations

A key strength of this study lies in its application of (GWPR) to examine spatial heterogeneity in substance-related mortality across Iran, enabling identification of region-specific risk factors that can inform localized intervention strategies. Additionally, the comprehensive coverage of multiple provinces provides valuable insights into nationwide patterns.

It is worth noting that, in our study, the GWPR model poorly predicted counties classified as high-high (H-H). This could be because the model makes predictions by considering the neighborhoods of each region, but in northwestern regions such as East Azerbaijan province, which has no neighbors, the model performed poorly.

Nevertheless, several limitations warrant consideration. First, the ecological study design precludes inferences about individual-level associations, as observed relationships at the county level may not reflect individual risk profiles. The cross-sectional nature of the data further limits our ability to establish temporal sequences or causal relationships between exposures and outcomes. Second, the social stigma surrounding addiction in Iranian society may lead families to conceal substance-related deaths, potentially resulting in underreporting and misclassification of mortality causes. Third, variations in the completeness and accuracy of death registration systems across provinces, particularly in remote or under-resourced areas, may have affected the reliability of mortality estimates in certain regions. Finally, our data are limited to the period up to 2018, and more recent data are needed to capture the potential effects of the COVID-19 pandemic on substance-related mortality patterns, as the pandemic has substantially altered socioeconomic conditions, healthcare access, and substance use behaviors.

## 5 Conclusions

Our study revealed that substance-related deaths across Iranian counties show significant spatial clustering, with high-risk areas concentrated in East Azerbaijan province. This pattern reflects the complex interplay of geographic location along substance trafficking routes, urbanization, and regional cultural contexts. However, apparent low-risk areas in southern regions may reflect data quality limitations rather than a true reduction in mortality. The predominance of deaths among men aged 30-39 years and variations by marital status underscore the multifaceted nature of risk factors. These findings can guide policymakers and health officials in developing targeted prevention strategies and resource allocation to address the burden of substance use disorders in high-risk communities.

## Acknowledgments

We sincerely thank the Iran Legal Medicine Organization (ILMO, Statistical Center of Iran for providing the valuable data used in this study. We also extend our gratitude to our colleagues for their collaboration and support. Additionally, we appreciate the insightful guidance of professional experts, particularly their consultants in variable adjustment, which greatly enhanced the quality of our research.

## Ethics Approval

The obtained ethical code for this was IR.SBMU.PHNS.REC.1398.107.

## Data Availability Statement

The data supporting the findings of this study are available [insert location, e.g., in a publicly accessible repository, upon reasonable request.

## Author Contributions

FB, AK, EK and SH conceptualized and supervised this project.

FB, EK, AR, SH, A H and EK contributed in methodology, investigation and validation.

FB carried out data curation and collection.

FB, AK and SH helped in formal analysis.

FB and YA contributed in visualization and software.

FB and FJ were major contributors in writing the original draft.

FB, AH, SA, SH, AR and EK critically reviewed and edited the manuscript.

All authors have read and approved the final version of the manuscript.

## Supporting Information Captions

Table S1: Description of variables included in the analysis

Table S2: The number and incidence rate of substance-related mortality in the provinces and counties of Iran per one million population

Fig S1: Spatial autocorrelation of substance-related mortality rate with Global Moran’s I

Fig S2: Spatial autocorrelation of substance-related mortality rate with Getis G

Table S3: Cluster and Outlier of Substance-related mortality rates in Iran (Local Moran’s I)

**Fig S1:**
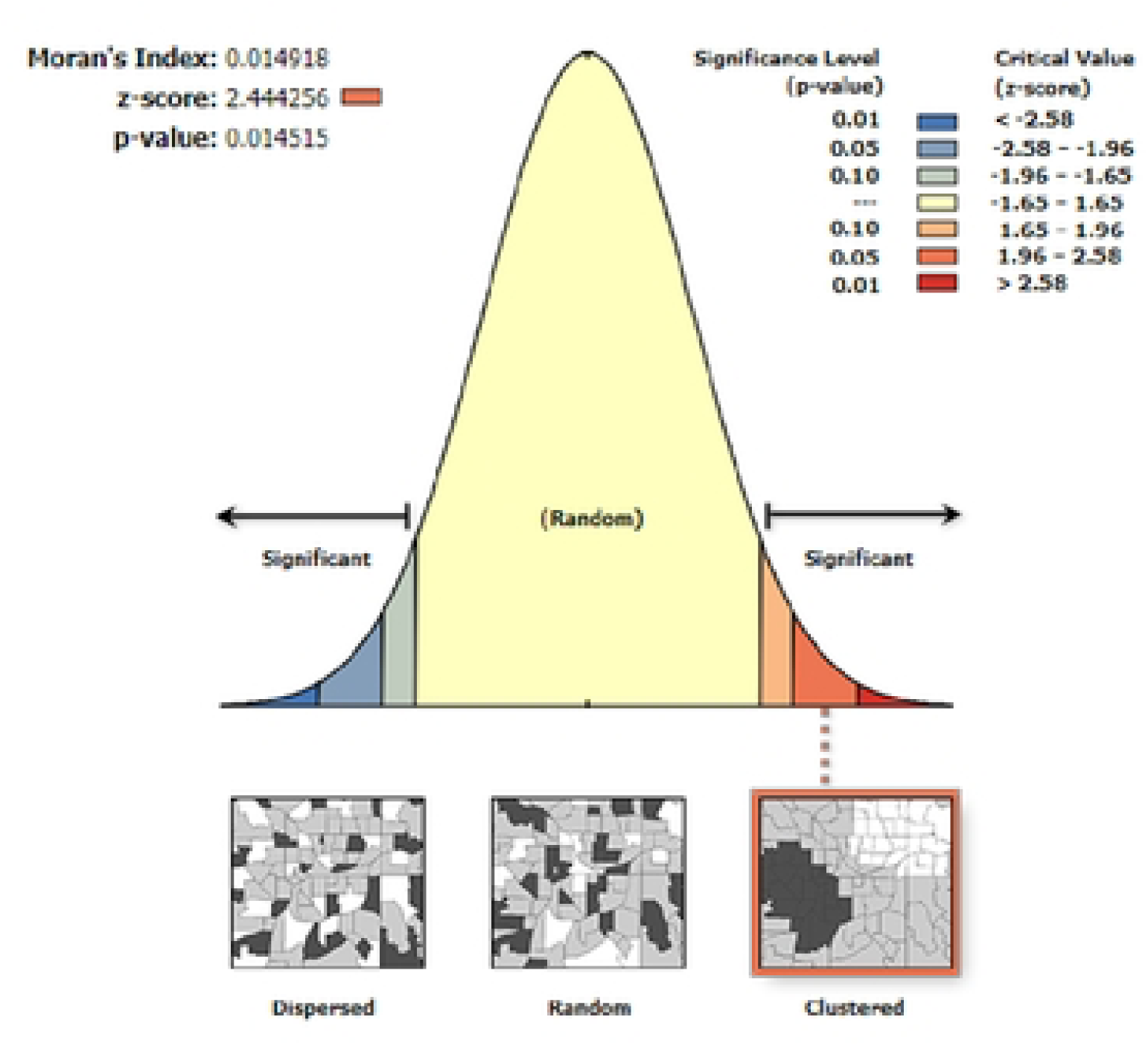
Spatial autocorrelation of substance-related mortality rate with Global Moran’s I

**Fig S2:**
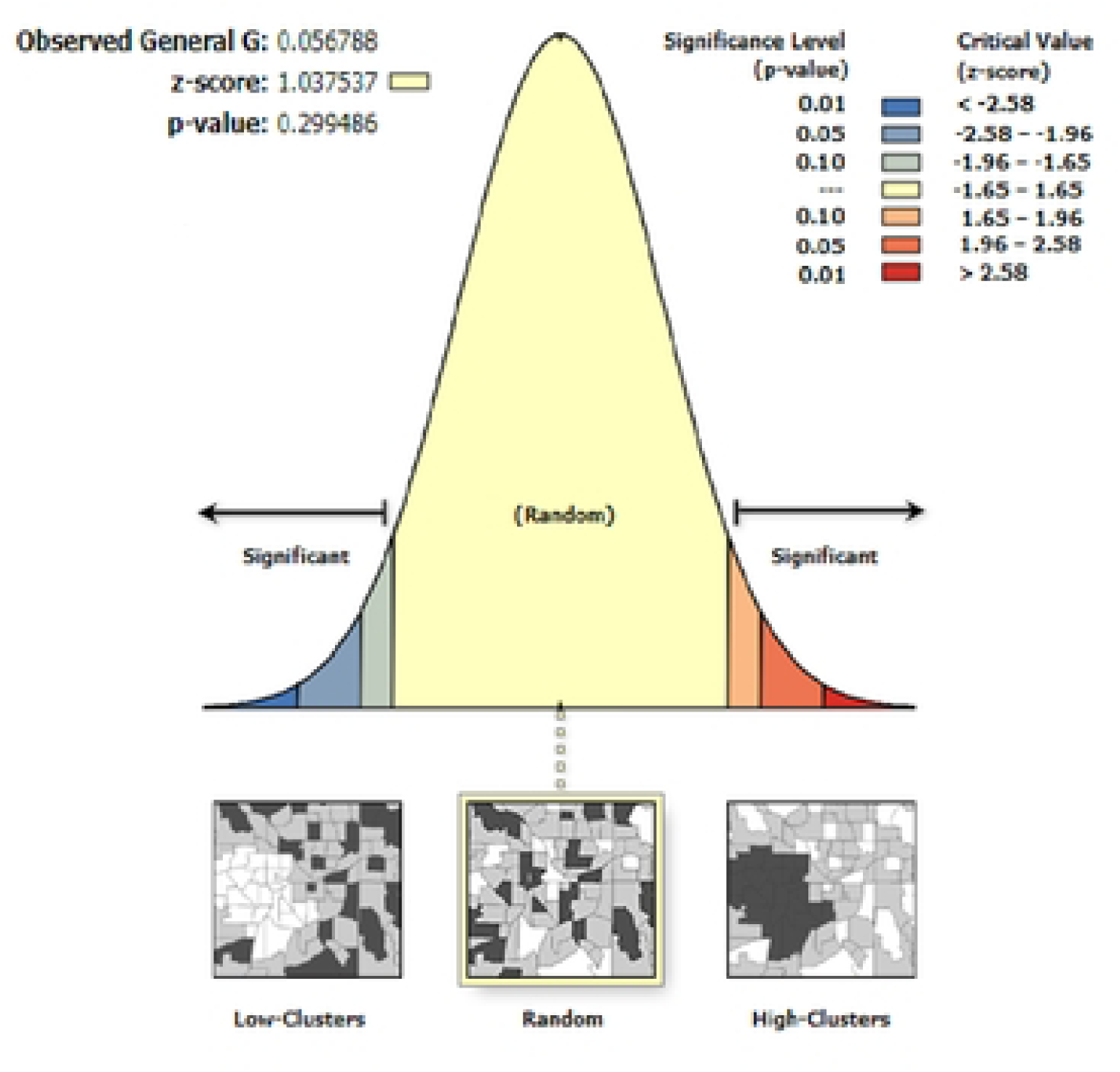
Spatial autocorrelation of substance-related mortality rate with Getis G

